# *CIZ1-*LOSS CAUSES FEMALE-SPECIFIC AUTOSOMAL NEURODEVELOPMENTAL DISORDER THROUGH DEFECTIVE X-INACTIVATION MAINTENANCE

**DOI:** 10.64898/2025.12.19.25342284

**Authors:** Thomas Besnard, Agnese Loda, Emma Kneuss, Laura Do Souto Ferreira, Frédéric Ebstein, Virginie Vignard, Wallid Deb, Sébastien Küry, Meriem M. Derradji-Costea, Gaëlle Landeau-Trottier, Patricia Talarmain, Eva Trochu, Valeria Dutan Patino, Amélie Piton, Jean-Baptiste Lamouche, Marc Gibaud, Jana Behunova, Franco Laccone, Hannes Steinkellner, Xavier Blanc, Martin Broly, Emmanuelle Ranza, Iman Abumansour, Mais O Hashem, Hanan E Shamseddin, Rayyan Albarakati, Majid Alfadhel, Gabriela Oprea, Aboulfazl Rad, Lama Abdullah Alabdi, Bader Alhaddad, Khadijah Bakur, Talal Alanazi, Jihan Madani, Ehsan Ghayoor Karimiani, Henry Houlden, Reza Maroofian, Gavin N McNee, Grant S Stewart, Stylianos Antonarakis, Fowzan Alkuraya, Stéphane Bézieau, Edith Heard, Benjamin Cogné, Bertrand Isidor

**Affiliations:** Nantes Université, CHU Nantes, Service de Génétique Médicale, Nantes, France; Nantes Université, CHU Nantes, CNRS, INSERM, l’institut du thorax, Nantes, France; European Molecular Biology Laboratory, Directors’ Research, 69117 Heidelberg, Germany; The Francis Crick Institute, London, UK; Institute for Genetics and Molecular and Cellular Biology (IGBMC), University of Strasbourg, CNRS UMR7104, INSERM U1258, Illkirch, France; Department of Pediatrics and Pediatric Emergency, Nantes University Hospital, Nantes, France; Institute of Medical Genetics, Center for Pathobiochemistry and Genetics, Medical University of Vienna, 1090, Vienna, Austria; Medigenome, Swiss Institute of Genomic Medicine, Geneva, Switzerland; Department of Molecular Genetics and Cytogenomics, CHU Montpellier, CeRéMAIA, IRMB, Univ Montpellier, INSERM, Montpellier, France; Medical Genetic Department, Umm Alqura University, Makkah; Department of Translational Genomics, Center for Genomics Medicine, King Faisal Specialist Hospital and Research Center, Riyadh, Saudi Arabia; Genetics and Precision Medicine department (GPM), King Abdullah Specialized Children’s Hospital (KASCH), King Abdulaziz Medical City, Ministry of National Guard Health Affairs (MNG-HA), Riyadh, Saudi Arabia; Medical Genomic Research Department, King Abdullah International Medical Research Center(KAIMRC), King Saud Bin Abdulaziz University for Health Sciences(KSAU-HS), Ministry of National Guard Health Affairs (MNG-HA), Riyadh, Saudi Arabia; King Salman Center for Disability Research, Riyadh, 11614, Saudi Arabia; Arcensus GmbH, Rostock, Germany; Lifera Omics, Riyadh, 13519, Saudi Arabia; Neuroimmunology, Neuroimmunology and Headache Department, Neuroscience center, King Faisal Specialist Hospital & Research Center, Riyadh, Saudi Arabia; College of Medicine, Alfaisal University, P.O. Box 50927, Riyadh, 11533, Saudi Arabia; Department of Neuromuscular Diseases, UCL Queen Square Institute of Neurology, London, UK; Department of Cancer and Genomic Sciences, University of Birmingham, Birmingham B15 2TT, UK; Department of Genetic Medicine, University of Geneva Medical School, Geneva, Switzerland; Laboratoire SeqOIA, Paris, France

**Keywords:** CIZ1, X-chromosome inactivation, XIST RNA, neurodevelopmental disorder, female-specific disease

## Abstract

We report an autosomal recessive neurodevelopmental disorder exclusively affecting females carrying biallelic loss-of-function variants in *CIZ1*, a gene encoding a nuclear matrix protein essential for the maintenance of X-chromosome inactivation. Eight unrelated affected females were identified, whereas male siblings with biallelic variants were asymptomatic. We showed that loss of CIZ1 compromises the maintenance of X-chromosome inactivation, leading to an abnormal overexpression of a subset of X-linked genes.

## MAIN

X-chromosome inactivation (XCI), initiated by the 17-kb long noncoding RNA X-inactive specific transcript (XIST), ensures dosage compensation between females (XX) and males (XY) by transcriptionally silencing the majority of genes on one X-chromosome in each female cell during embryonic development (Penny et al., 1996; Galupa & Heard, 2018; Patrat et al., 2020; Loda et al., 2022). Disruption of XCI stability has been implicated in human disorders, particularly in severe neurodevelopmental impairment observed in females with X ring chromosome lacking XIST expression (Migeon et al., 1993). However, human monogenic disorders directly caused by defective XCI maintenance have not previously been reported.

We investigated a family with a young girl presenting severe neurodevelopmental delay (F1-II-4) (**Fig. 1-A**). Trio genome sequencing of the proband and her parents was performed as part of the French National Genomics plan (PFMG2025; (PFMG2025 contributors, 2025)). Although no pathogenic variants were identified in known OMIM genes, we detected compound heterozygosity of two loss-of-function (LOF) variants in *CIZ1* (MANE transcript NM_001131016.2; 9q34.11): an intragenic deletion (c.1944-2806_2296-757del; deletion of exons 12 to 14) on the maternal allele and a stop-gain variant (c.2293C>T; p.(Gln765Ter)) on the paternal allele.

**Figure 1:**
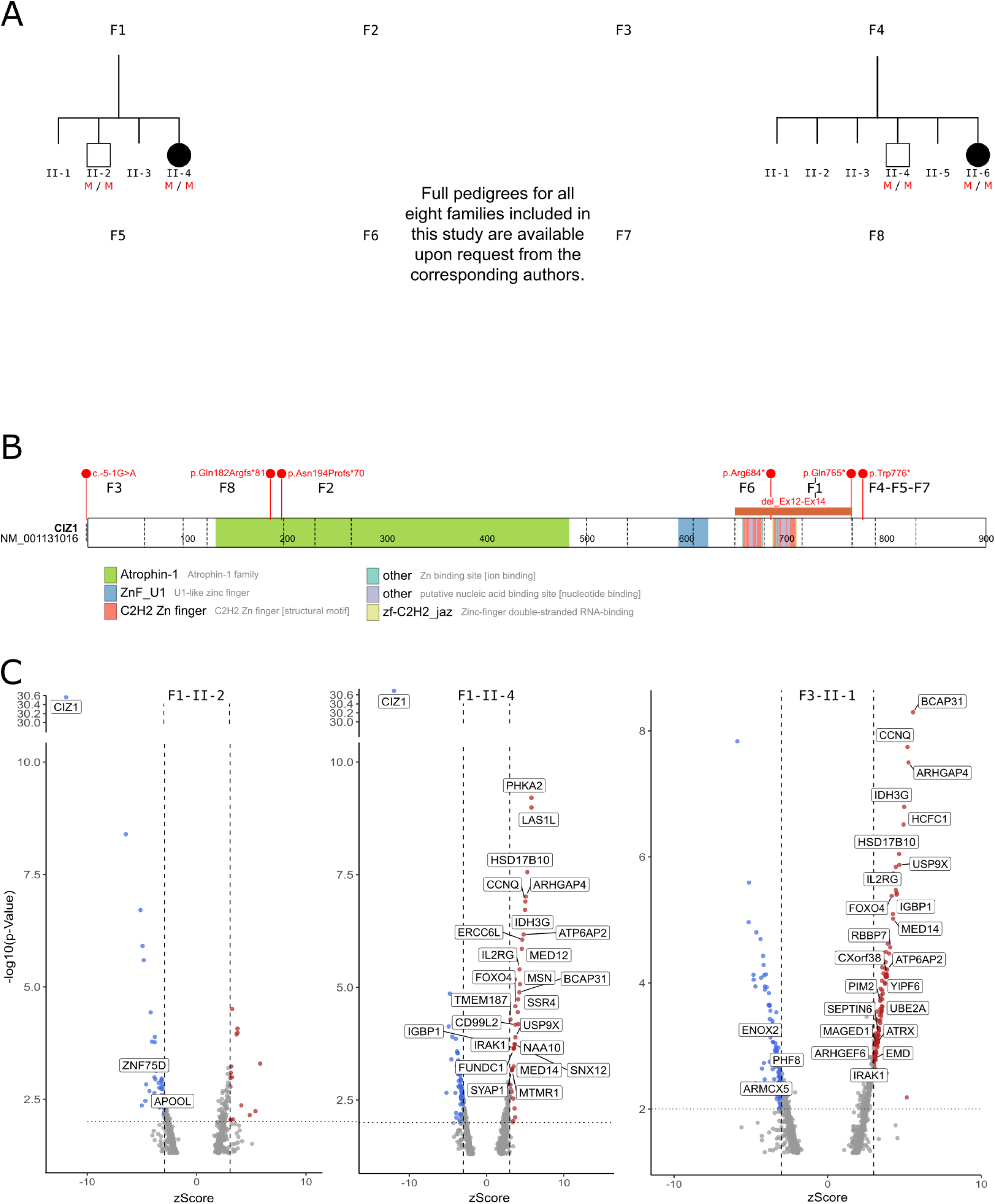
CIZ1 Mutations: Pedigree, Gene Mapping, and Expression profiling. **A-Pedigrees of eight families (F1–F8) carrying *CIZ1* variants.** Black symbols indicate individuals clinically affected by the neurodevelopmental disorder. Dashes indicate wild-type *CIZ1* alleles, red “M” for mutated *CIZ1* alleles, and the question marks represent unknown status. **B-Schematic representation of *CIZ1* (NM_001131016) adapted from ProteinPaint (St. Jude Children’s Research Hospital).** Red circles indicate the positions of variants found in affected individuals from families F1–F8. The orange rectangle represents the deletion spanning exons 12 to 14. **C-Volcano plots** showing significantly differentially expressed genes in individuals F1-II-2 (healthy compound heterozygous male sibling), F1-II-4, and F3-II-1 (both affected females). The x-axis represents the z-score, and the y-axis shows the statistical significance as -log10(p-value). Red and blue points represent significantly upregulated and downregulated genes, respectively. Annotated genes include *CIZ1* and those significantly differentially expressed from chromosome X.

Through GeneMatcher, we identified seven additional unrelated individuals, all females, with a similar phenotype that included severe language impairment (6/6), hypotonia (8/8), early-onset global and severe developmental delay (6/7), and seizures (4/8) (**Table 1, Supp. Table 1**). All seven variants identified by exome/genome sequencing in *CIZ1* in these patients are putative LOF and absent in gnomAD (v.4.1.0) (**Fig. 1-B**). A recurrent c.2328G>A variant was identified in three unrelated Middle East families (families F4, F5, and F7) on the same haplotype background, confirming a founder effect. For all available parental samples, segregation analyses showed that unaffected parents were heterozygous carriers (**Fig. 1-A**).

**Table 1.**
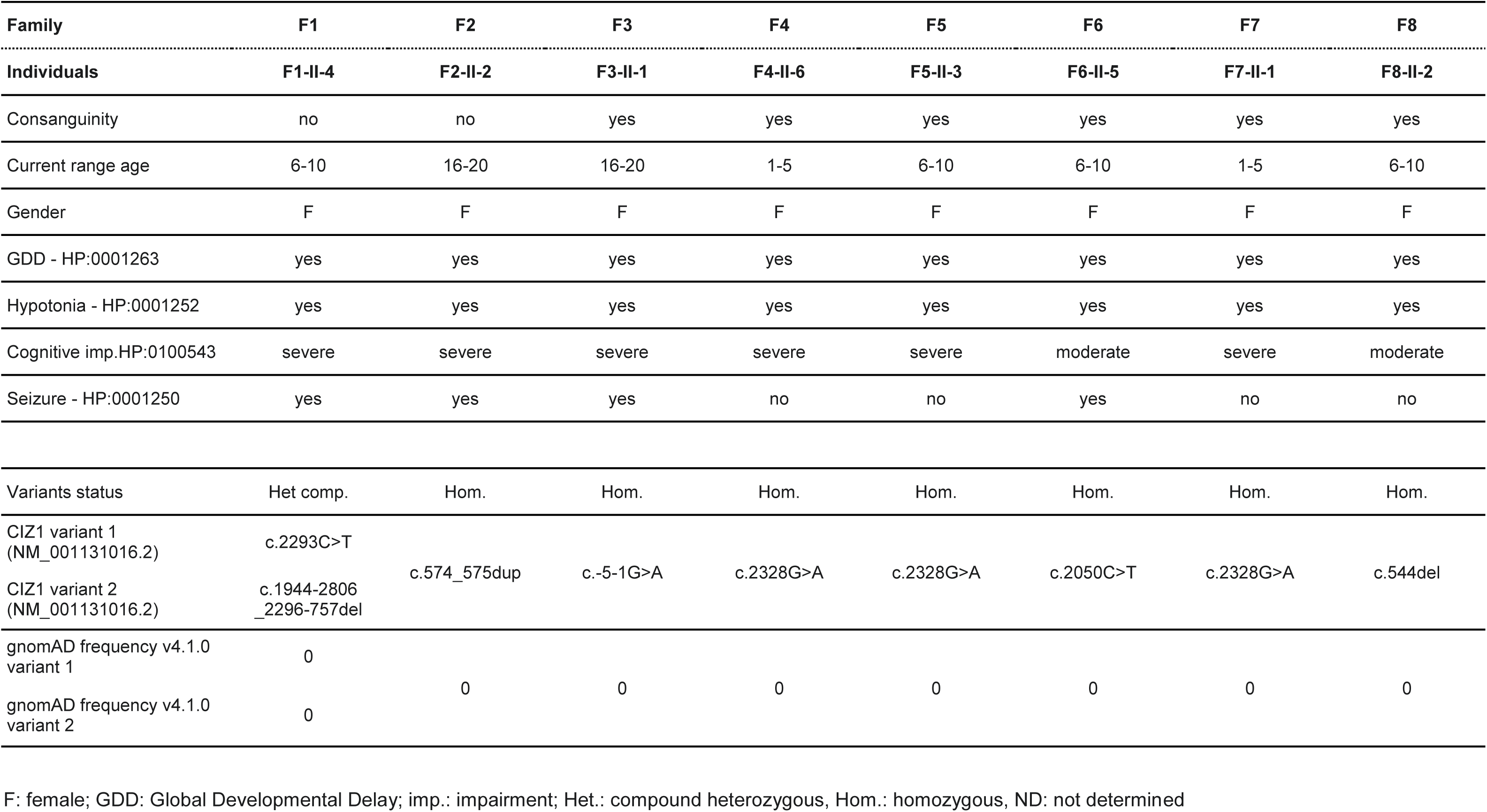
Simplified clinical table

Segregation analysis also identified two asymptomatic male siblings (F1-II-2 and F4-II-4) carrying the same biallelic *CIZ1* variants as their affected female sibling. To investigate the absence of clinical signs in males, we performed RNA-Seq on short-term lymphocyte cultures from two affected females (F1-II-4 and F3-II-1) and one unaffected male (F1-II-2), all carrying biallelic loss-of-function variants in *CIZ1*. We used OUTRIDER to identify gene expression outliers relative to a control cohort of 155 unrelated individuals. *CIZ1* expression was similarly reduced in F1-II-4 and her healthy male sibling F1-II-2 (log2 fold change of -3.79 and -3.37). In contrast, for F3-II-1, who carries the homozygous c.-5-1G>A variant, *CIZ1* expression was not decreased (**Fig. 1-C**), but an alternative 3’ splice site was used, abolishing the canonical start codon (**Supp. Fig 1-A**). Reduced CIZ1 protein levels were confirmed by Western blot analysis of lymphocyte lysates (**Supp. Fig 1-B**).

We next examined additional differentially expressed genes (adjusted p-value < 0.05). Remarkably, almost all expression outliers in affected females were upregulated genes located on the X-chromosome (8/8 and 6/7 in F1-II-4 and F3-II-1, respectively), whereas the unaffected male exhibited only 3 autosomal outliers (**Supp. Table 2**). Using a less stringent threshold (p-value < 0.01) to assess the overlap between the three individuals (**Supp. Fig 2-A**), we observed few common dysregulated genes in the siblings of family F1 (F1-II-4 and F1-II-2); however, we identified 27 upregulated genes in the two affected females F1-II-4 and F3-II-1. The vast majority (25/27) of these shared outliers mapped to the X-chromosome. Several of these genes have previously been involved in neurodevelopmental disorders: *SLC35A2* (Kodera et al., 2013), *USP9X* (Reijnders et al., 2016), *BCAP31* (Whalen et al., 2021), *SSR4* (Losfeld et al., 2014), *OFD1* (Budny et al., 2006), *ATP6AP2* (Gupta et al., 2015), *HSD17B10* (Yang et al., 2009), *LAS1L* (Hu et al., 2016).

CIZ1 binds to the E-repeat of XIST RNA (Dixon-McDougall & Brown, 2022; Sunwoo et al., 2017) and is required to anchor XIST to the inactive X (Xi) territory, thereby preventing its diffusion throughout the nucleoplasm. *CIZ1* knockout in cells leads to the delocalization of Xist RNA and the upregulation of a subset of X-linked genes, suggesting a direct role for CIZ1 in XCI maintenance through control of Xist localization (Ridings-Figueroa et al., 2017; Jacobson et al., 2022; Sofi & Coverley, 2023). Thus, we hypothesized that impaired maintenance of XCI could explain why the disorder would manifest exclusively in females.

In order to identify genes escaping XCI in lymphocyte cultures, we determined the X-linked genes upregulated in control females compared to a male-only cohort. We identified 24 significantly upregulated genes (**Supp. Fig 2-B**): 17/24 (red genes, **Supp. Fig 2-B**), including *SMC1A*, *KDM5C,* and *KDM6A*, which are known constitutive escapees (escape genes) that resist XIST-mediated silencing from the onset of XCI and are expressed from both the active (Xa) and inactive (Xi) X-chromosomes in most cell types and individuals (Tukiainen et al., 2017; Garieri et al., 2018). The remaining seven genes (7/24) are classified as inactive by Tukiainen et al. (Tukiainen et al., 2017) We then compared the 25 upregulated genes (**Fig. 2-A**) in both *CIZ1* LOF females to the 24 escapees highlighted in lymphocyte cultures (**Supp. Fig 2-B**). Eleven genes (11/25) overlapped (**Supp. Fig 2-C**), indicating that some genes already expressed by Xi (escaping XCI) in control females were even more expressed in *CIZ1* LOF females. The remaining 14/25 (56%) were not significantly upregulated in the female control cohorts. Mapping these 25 genes along the X-chromosome (**Fig. 2-A**; **Supp. Fig 2-C**) showed that 13 of the 14 non-escape outliers lie in the same genomic regions that also harbor physiological escapees, defining four clusters: two positioned on either side of the centromere, one on the short arm (Xp22.12p22.2), and one in the subtelomeric Xq region (**Fig 2-A**). As CIZ1 is known to anchor XIST to the nuclear matrix (Ridings-Figueroa et al., 2017), we performed XIST RNA fluorescence *in situ* hybridization analysis (FISH) in fibroblasts derived from the affected female in family 1 (F1-II-4) to assess XIST RNA cellular localization in these individuals. Patient fibroblasts displayed a diffuse XIST RNA localization pattern in contrast to control fibroblasts, which exhibited a single compact XIST cellular distribution corresponding to the Xi (**Fig. 2-A**). These data are consistent with previous studies on *Ciz1*-null mouse fibroblasts, which also showed impaired Xist localization to the Xi nuclear territory and upregulation of a subset of X-linked genes (Ridings-Figueroa et al., 2017; Sofi & Coverley, 2023). CIZ1 is a ubiquitous nuclear matrix protein initially characterized for its role in the initiation of DNA replication and more recently for anchoring XIST to the nuclear matrix through binding to its repeat E region (Sofi & Coverley, 2023). During XCI initiation, XIST RNA spreads in *cis* and coats one of the two X-chromosomes, triggering transcriptional silencing of nearly all X-linked genes. However, a subset of genes resist XIST-mediated silencing and completely or partially escape XCI, thus remaining expressed from the otherwise silent Xi (Tukiainen et al., 2017). These escape genes are typically expressed at higher levels in females than in males and are expected to contribute to sex differences in contexts such as sex-biased disease susceptibility, as observed in autoimmunity (Fang et al., 2021). Loss of XIST can lead to the upregulation of escapees or reactivation of silenced oncogenes, hereby causing cancer development (Yildirim et al., 2013; Ridings-Figueroa et al., 2017; Richart et al., 2022). Xist RNA has also been shown to modulate expression levels of escape and variably escaping genes in differentiated cells (Yildirim et al., 2013; Richart et al., 2022; Hauth et al., 2024).

**Figure 2:**
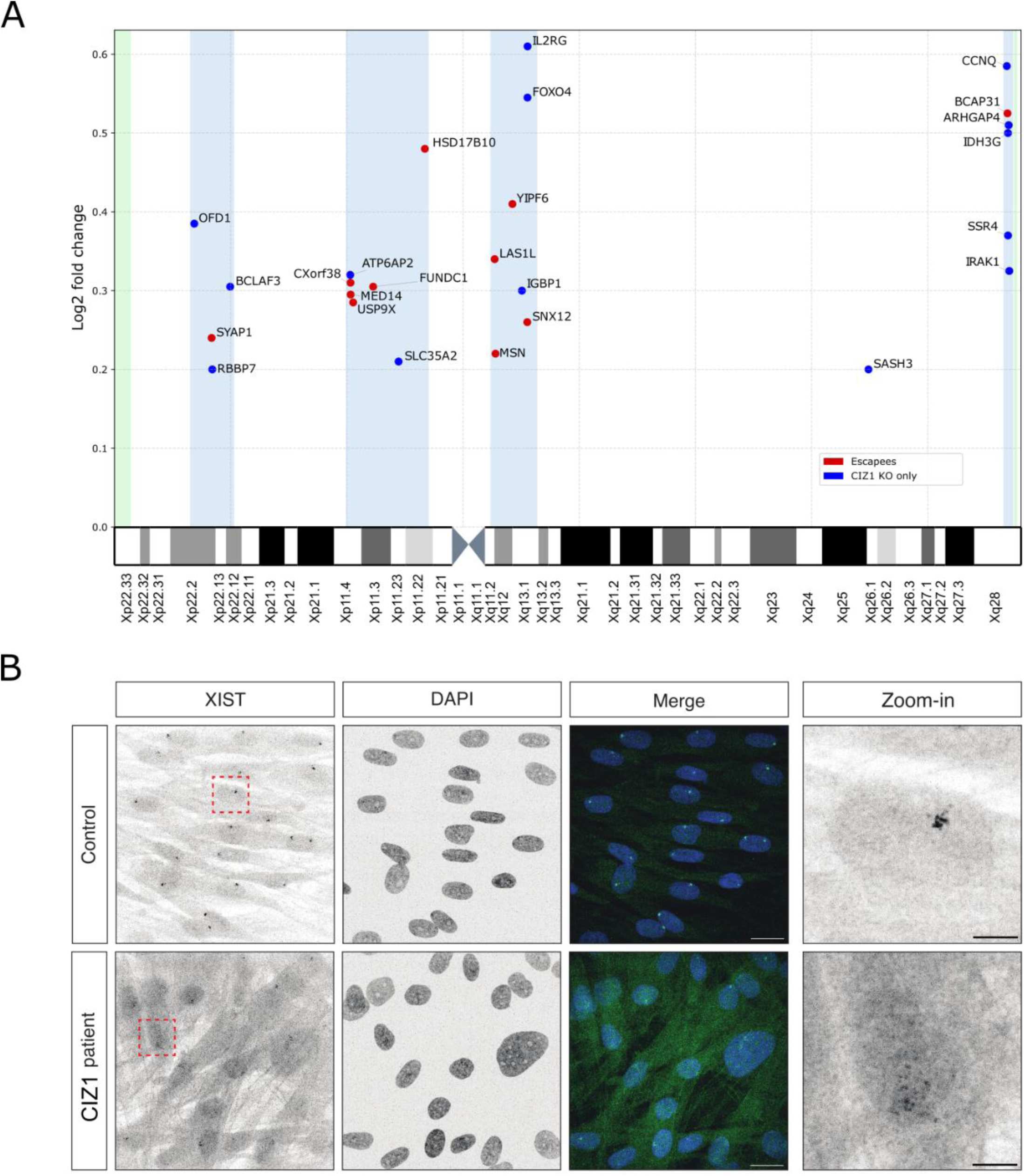
Consequences of *CIZ1* biallelic loss-of-function. **A-Distribution of overexpressed genes along the X-chromosome in lymphocyte cultures from girls with *CIZ1* biallelic loss-of-function variants.** The plot shows the median log2 fold change of significantly overexpressed genes (OUTRIDER; p-value < 0.01) shared by two girls with biallelic *CIZ1* loss-of-function compared to 155 controls. Chromosomal position is indicated on the x-axis. Red dots indicate genes identified as escapees in our lymphocytes culture protocol while blue dots correspond to genes not overexpressed in our control cohort females compared to a male-only cohort (see **Supp Figure 2-B & 2-C**). **B-XIST RNA Delocalization in Patient Fibroblasts. XIST RNA Fluorescence *in situ* Hybridization (RNA FISH) analysis shows the localization of the XIST RNA.** The control (top row) displays a condensed XIST domain, characteristic of the Xi territory. CIZ1 patient (bottom row) exhibits a diffuse XIST signal suggesting CIZ1 is essential for anchoring XIST in the nuclear matrix. (green) Xist; (blue) DAPI/DNA. Scale bar for Merge: 20µm; scale bar for zoom-in: 5µm

Here we show that CIZ1 is required for correct localization of XIST to the Xi territory in fibroblasts derived from *CIZ1*-deficient females and that XIST delocalization throughout the nucleoplasm results in upregulation of escape genes and their neighbouring X-linked genes in lymphocytes. Our findings suggest that CIZ1 is necessary for proper *XIST*-dependent regulation of escapees in somatic cells, and can disrupt the expression of topologically associated genes along the X-chromosome.

In mice, *Ciz1* knock-out causes a female-specific lymphoproliferative disorder (Ridings-Figueroa et al., 2017). Here, we show that in humans biallelic *CIZ1* deficiency results in a female-specific neurodevelopmental impairment. A neurodevelopmental phenotype also occurs in females with ring X-chromosomes or XIST deletions, in whom X-linked gene expression is upregulated (Migeon et al., 1993). Together, these observations support a model in which loss of XCI maintenance leads to de-dampening of escapee expression levels or reactivation of a limited number of genes leading to neurodevelopmental disease in females. More extensively, our findings lay the foundations for a new paradigm establishing the contribution of XCI maintenance to normal female neurodevelopment and suggest that female-specific neurodevelopmental disorders may arise from pathogenic variants in autosomal genes encoding regulators of XCI.

## CLINICAL DATA

Detailed clinical and genealogical information for all eight families included in this study is available upon request from the corresponding authors.

### Family 1

Compound heterozygous *CIZ1* variants (c.1944-2806_2296-757del and c.2293C>T; p.Gln765* nonsense mutation); individual presented with early-onset severe developmental delay, severe language impairment, myoclonic epilepsy, and generalized hypotonia. An asymptomatic male relative carried the same biallelic variants.

### Family 2

Homozygous *CIZ1* frameshift variant (c.574_575dupAC; p.Asn194Profs*70); individual presented with severe developmental delay, severe language impairment, hypotonia, and early-onset seizures. Additional features noted including orthopedic abnormalities and growth variation.

### Family 3

Homozygous *CIZ1* splice site variant (c.-5-1G>A; p.?); individual presented with severe developmental delay, severe language impairment, myoclonic epilepsy, and hypotonia. Born to consanguineous parents. Variable dysmorphic features noted.

### Family 4

Homozygous *CIZ1* nonsense variant (c.2328G>A; p.Trp776*); individual presented with severe developmental delay, severe language impairment, hypotonia, and seizures. An asymptomatic male relative carried the same biallelic variant. Consanguineous family. This recurrent variant was identified in Families 5 and 6 on identical haplotype.

### Family 5

Homozygous recurrent *CIZ1* nonsense variant (c.2328G>A; p.Trp776*); individual presented with severe developmental delay, severe language impairment, hypotonia, and ataxia. Preterm birth with low birth weight documented. Consanguineous family background.

### Family 6

Homozygous *CIZ1* nonsense variant (c.2050C>T; p.Arg684*); individual presented with severe developmental delay, hypotonia, and infantile spasms requiring medical management. Consanguineous family. Growth restriction and subtle dysmorphic features documented.

### Family 7

Homozygous recurrent *CIZ1* nonsense variant (c.2328G>A; p.Trp776*); individual presented with severe developmental delay, severe language impairment, hypotonia, and growth restriction. A de novo variant in SMARCA2 (c.620A>G; p.Gln207Arg) was also identified. Consanguineous parents.

### Family 8

Homozygous *CIZ1* frameshift variant (c.544del; p.Gln182Argfs*81); individual presented with moderate developmental delay, language impairment, and persistent hypotonia. Consanguineous family background. A healthy male relative was identified.

## MATERIALS AND METHODS

### Study participants

Individuals were recruited *via* an international, multi-center collaboration with research and diagnostic sequencing laboratories and medical genetics departments. Individuals were clinically evaluated in separate centers and initially enrolled in different centers to investigate the molecular basis of their developmental disorders: Nantes University Hospital (Nantes, France), Medical University of Vienna (Vienna, Austria), Medigenome Swiss Institute of Genomic Medicine (Geneva, Switzerland), King Faisal Specialist Hospital and Research Center (Riyadh, Saudi Arabia), Children’s Hospital of Philadelphia (Pennsylvania, United States), and University College London (London, Great Britain). Contact between researchers was facilitated by use of the web-based tool GeneMatcher (Sobreira *et al*., 2015). Inclusion criteria were rare biallelic variants in CIZ1 carried by individuals with compatible phenotypes. Photographs and clinical data reports were provided by each attending physician and/or a detailed review of medical records.

### Ethics statement

Informed consent to participate in the study was obtained from each individual’s legal representative, in accordance with local institutional review boards (Comité consultatif sur le traitement de l’information en matière de recherche; number CCTIRS: 14.556). Samples and clinical information were obtained for each patient following local procedures and ethical standards. When needed, additional written consent to use photographs in this publication was also obtained.

### Sequencing and bioinformatics analysis

Whole exome/genome sequencing (WES/WGS) was performed on blood-derived DNA samples using each center’s analysis platforms and Next Generation Sequencing (NGS) pipeline. Exome data were interpreted in agreement with local practices. cDNA and protein sequence variants are described in accordance with the recommendations of the Human Genome Variation Society using the following MANE Select transcript: NM_001131016.2 (RefSeq)/ ENST00000372938.10 (Ensembl) as the reference sequence, and genomic coordinates from GRCh38. Priority was given to rare exonic or donor/acceptor splicing variants in accordance with the pedigree and phenotype, fitting a recessive (homozygous or compound heterozygous) model and/or variants in genes previously linked to developmental delay, intellectual disability, and other neurological disorders. Upon whole-exome sequencing analysis, *CIZ1* variants were selected and all the other candidate variants were further verified through the use of Sanger sequencing.

### RNA-seq from Lymphocytes short term culture

#### Blood cells Isolation, Culture, and RNA Extraction

Peripheral blood mononuclear cells (PBMCs) were isolated from EDTA-anticoagulated blood samples (2-4 ml, collected within 48 hours) using UNISEP+ tubes (EUROBIO), according to the manufacturer’s protocol. Cells were cultured in 6-well plates (0.5-2.0×10^6 cells/well) in lymphocyte-stimulating medium (Chromosome Medium P, EUROCLONE) for 48-72 hours at 37°C with 5% CO2 (4 wells). RNA was then extracted from one dry pellet of each condition, using the NucleoSpin RNA Plus kit (MACHEREY-NAGEL) following the manufacturer’s protocol.

#### Library RNA preparation and sequencing

Stranded RNA-Seq libraries were prepared from 100ng of total RNA using the SureSelect XT-HS2 kit (Human All Exon V8 capture probes, Agilent) on the automated Magnis NGS Prep System (Agilent). Pre-capture and post-capture PCR amplifications were performed with 12 and 10 cycles, respectively. RNA sequencing was performed on Illumina’s NextSeq 550 (HighOutput 2×75bp) to obtain 25-30 million paired-end reads per sample (16 samples per run).

#### RNA-seq OUTRIDER and other analyses

Quality control of the FASTQ files was performed using FastQC (v.0.11.3). The reads were then cleaned based on base quality using the trimming tool fastp (v.0.23.4). Alignment was carried out with STAR (v2.7.11a) against the Ensembl GRCh38 (p13) reference genome using the corresponding GTF annotation file (v106). Gene-level counts were generated using HTSeq (v2.0.5), and expression outliers were subsequently identified with the R package OUTRIDER (v1.24).

The three CIZ1 samples were analyzed together with 155 probands with NDD and without CIZ1 biallelic variants. Escapees were evaluated on 10 females, each female was tested with a male cohort of 70 individuals. We retained escapees as genes upregulated in at least 5 females with a p-value < 0.01.

### XIST RNA fluorescence in situ hybridization (FISH)

RNA FISH on fibroblasts was performed as previously described (Chaumeil et al., 2008; Ranisavljevic et al., 2017). Fibroblasts were grown on fibronectin-coated coverslips. Cells were fixed with 3% formaldehyde in PBS for 10 min at room temperature and washed three times with PBS. Cells were permeabilized with an ice-cold permeabilization buffer (1X-PBS, 0.5% Triton X-100, 2 mM vanadyl-ribonucleoside complex (New England Biolabs)) for 4 min on ice. Coverslips were stored in 70% ethanol at -20 °C. Cells were dehydrated in increasing ethanol concentrations (80%, 95%, 100%) and quickly air dried. Probes were prepared from the VI.34 Flp-In T-Rex XIST cDNA DH10B construct (Chow et al., 2007) and fluorescently labelled by nick translation (Abbott) using Cy5-labeled dUTP (Merck). Labelled probes were co-precipitated with mouse Cot-1 DNA (Thermo Fisher) in the presence of ethanol and salt, resuspended in formamide, denatured at 75 °C for 8 min and competed at 37 °C for 40 min. Probes were hybridised in FISH hybridization buffer (50% formamide, 20% dextran sulfate, 2X SSC, 1 μg/μL BSA (New England Biolabs), 10 mM vanadyl-ribonucleoside complex) at 37 °C overnight. The next day coverslips were washed three times for 5 min with 50% formamide in 2X SSC at 42 °C, and three times for 5 min with 2X SSC at room temperature. DAPI (0.2 mg/mL) was added during the second wash and coverslips were mounted with ProLongTM Diamond antifade mountant (Thermo Fisher Scientific). Images were acquired using an OLYMPUS iXplore Spin-SR Spinning Disk microscope with a 100x objective. Images were analysed with ImageJ software (Fiji).

### SDS-PAGE and western blotting

T cells were expanded from peripheral blood mononuclear cells (PBMCs) isolated from whole blood using the Fonteneau protocol (PMID: 11684128). Cells were lysed in RIPA buffer (50 mM Tris-HCl pH 7.5, 150 mM NaCl, 2 mM EDTA, 1 mM N-ethylmaleimide, 10 µM MG-132, 1% NP-40, 0.1% SDS), and total protein concentration was determined using a bicinchoninic acid (BCA) assay (Thermo Fisher Scientific). Forty micrograms of protein per sample were separated on 4–12% SDS-PAGE gels (70 V, 3 h) and transferred onto PVDF membranes (15 V, 7 min). Membranes were blocked with 3% BSA in TBS containing 0.1% Tween-20 at room temperature and incubated overnight at 4 °C with primary antibodies against CIZ1 (31683-1-AP, Thermo Fisher Scientific) and GAPDH (clone 14C10, OZYME), with gentle agitation. After three washes in TBST (0.2% Tween-20), membranes were incubated for 1 h at room temperature with an HRP-conjugated anti-rabbit secondary antibody (7074S, OZYME). Protein detection was performed using an enhanced chemiluminescence (ECL) kit (Bio-Rad). Densitometric analysis of immunoreactive bands was performed using Fuji (ImageJ), an open-source image processing software.

## DATA AND CODE AVAILABILITY

All raw data presented in this paper are available to qualified researchers upon request.

## Data Availability

All data produced in the present study are available upon reasonable request to the authors

## ACKNOWLEDGMENTS

This work was carried out within the framework of the FHU GenOMedS with the support of the Health Cooperation Group of University Hospitals of the Great West (GCS HUGO). This work was also supported by the Groupama Foundation. G.S. and G.MN. received funding from the Medical Research Council (MRC) project grant (UKRI577). M.A, R.A., received funding from the King Salman Center for Disability Research through Excellence Research Center (No. KSCDR-ERC-2024-02).

## AUTHOR CONTRIBUTIONS

T.B., B.C. and B.I. conceived the study, developed the methodology, performed the investigation and wrote the original draft. A.L., E.K., E.H., contributed to conceptualization and wrote the original draft; M.B., L.DSF., W.D., S.K., J.B., H.S., F.A., G.S., S.B. and S.A. contributed to writing; L.DSF., E.K., F.E., V.V., A.P., J-B.L., P.T., V.DP., G.L-T., E.T., M.D-C. and G.MN. contributed to the investigation and performed analyses. M.B., M.G., E.R., X.B., H.E.S., B.A., I.A., M.H., L.A., J.M., T.A., K.B., F.L., A.R., G.O., M.A., R.A., R.M., E.G.K., H.H., J.B., and F.A. provided clinical information and resources. All authors read, reviewed and approved the manuscript, figures and data.

## WEB RESOURCES

GenBank: http://www.ncbi.nlm.nih.gov/genbank/

gnomAD: http://gnomad.broadinstitute.org/

GTEx: https://www.gtexportal.org/home/

Human Protein Atlas: https://www.proteinatlas.org/

Mobidetails: https://mobidetails.chu-montpellier.fr/

OMIM: http://www.omim.org/

UniProt: http://www.uniprot.org/uniprot/

UCSC Genome Browser: https://genome.ucsc.edu/

