## Supplementary Material for "*CIZ1-*LOSS CAUSES FEMALE-SPECIFIC AUTOSOMAL NEURODEVELOPMENTAL DISORDER THROUGH DEFECTIVE X-INACTIVATION MAINTENANCE"

**Supp. Fig 1:**

**A- IGV screenshot of RNA sequencing data for *CIZ1* locus in two affected individuals and controls.** The red box highlights the F1 compound heterozygous *CIZ1* individuals including the affected individual and the unaffected sibling, both exhibiting drastically reduced *CIZ1* transcript levels compared to other heterozygous relatives and unrelated controls. Bottomtrack corresponds to the individual with the homozygous c.-5-1G>A variant. The zoomed box on the bottom-right illustrates the start codon loss variant (“Startloss”) in this individual, contrasted with the unrelated controls.

**B- CIZ1 protein expression profile in T cells from healthy donors and individual F3-II-1.** RIPA lysates from T cells of two healthy donors (HD) and the patient were separated on 4-12% SDS-PAGE and analyzed by Western blotting using antibodies against CIZ1 and GAPDH (loading control). Asterisks (*) indicate non-specific bands. A representative experiment is shown. The right panel displays densitometric quantification of CIZ1 (≈140 kDa) immunoreactive bands, normalized to GAPDH and presented as the CIZ1/GAPDH ratio for HD and patient samples. Statistical analysis were performed using the Mann-Whitney test; *$p < 0.05$ was considered significant.

**Supp. Fig 1**

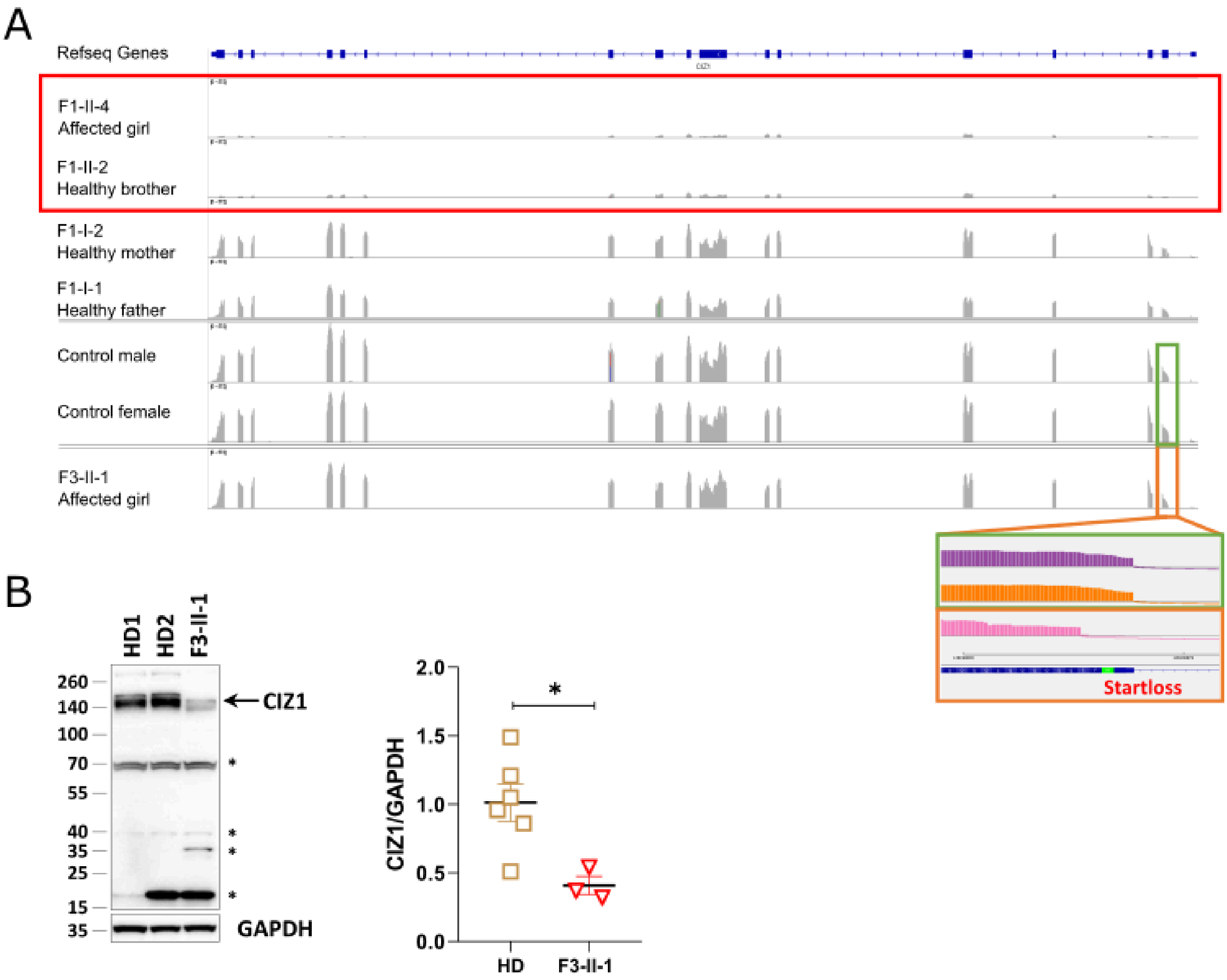
A
Refseq Genes
F1-II-4
Affected girl
F1-II-2
Healthy brother
F1-I-2
Healthy mother
F1-I-1
Healthy father
Control male
Control female
F3-II-1
Affected girl
Startloss
B
HD1
HD2
F3-II-1
260
140
100
70
55
40
35
25
15
35
CIZ1
GAPDH
*
CIZ1/GAPDH
2.0
1.5
1.0
0.5
0.0
HD
F3-II-1

**Supp. Fig. 2: Identification of X-inactivation escape genes and dysregulated genes in females with biallelic LOF *CIZ1* mutation.**

**A – Venn diagrams** showing genes significantly dysregulated in all chromosomes (p-value < 0.01, zScore < 0 for downregulated genes on the left; p-value < 0.01, zScore > 0 for upregulated genes on the right) across three individuals with biallelic *CIZ1* loss-of-function: two affected girls (F3-II-1, F1-II-4) and one unaffected male relative (F1-II-2, brother of F1-II-4).

**B – XCI escapees identified** in ten control females compared to 70 control males. Genes with p-value < 0.01 in at least 5 female samples were classified as X-inactivation escapees. The plot displays the median log2 fold change of these genes. Red dots correspond to genes previously described as escapees and black dots represent genes classified as inactive by Tukiainen (Tukiainen et al., 2017). Chromosomal position is shown on the x-axis.

**C – Venn diagram** comparing upregulated genes (OUTRIDER; p-value < 0.01) specific to CIZ1 female probands (yellow) versus those upregulated in control girls (green). Gene color coding is based on Tukiainen et al. (2017): red indicates genes previously described as escapees, grey represents constitutively inactive genes, blue denotes genes with variable XCI status, and green indicates genes not assessed in the original study.

**Supp. Fig. 2**

A

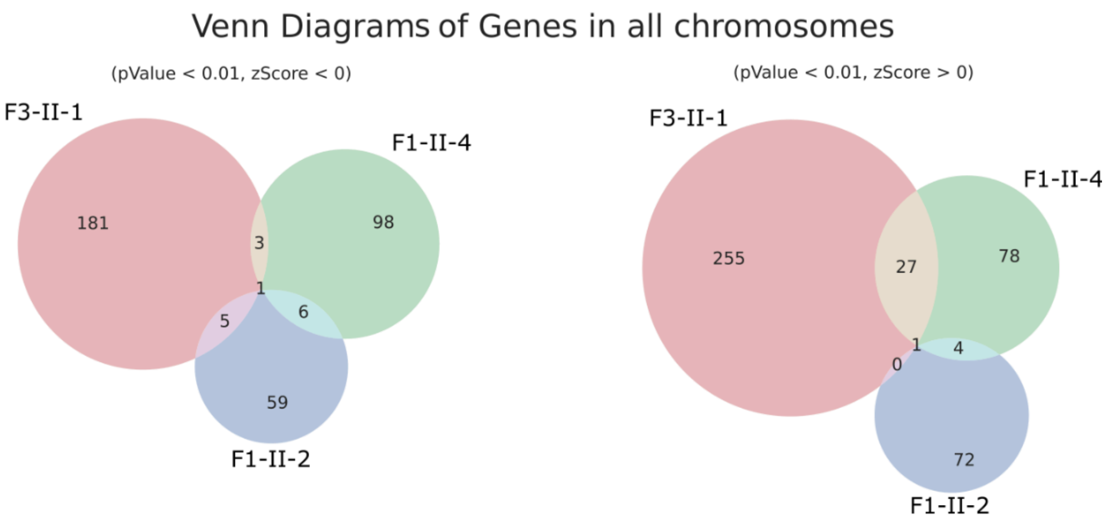


B

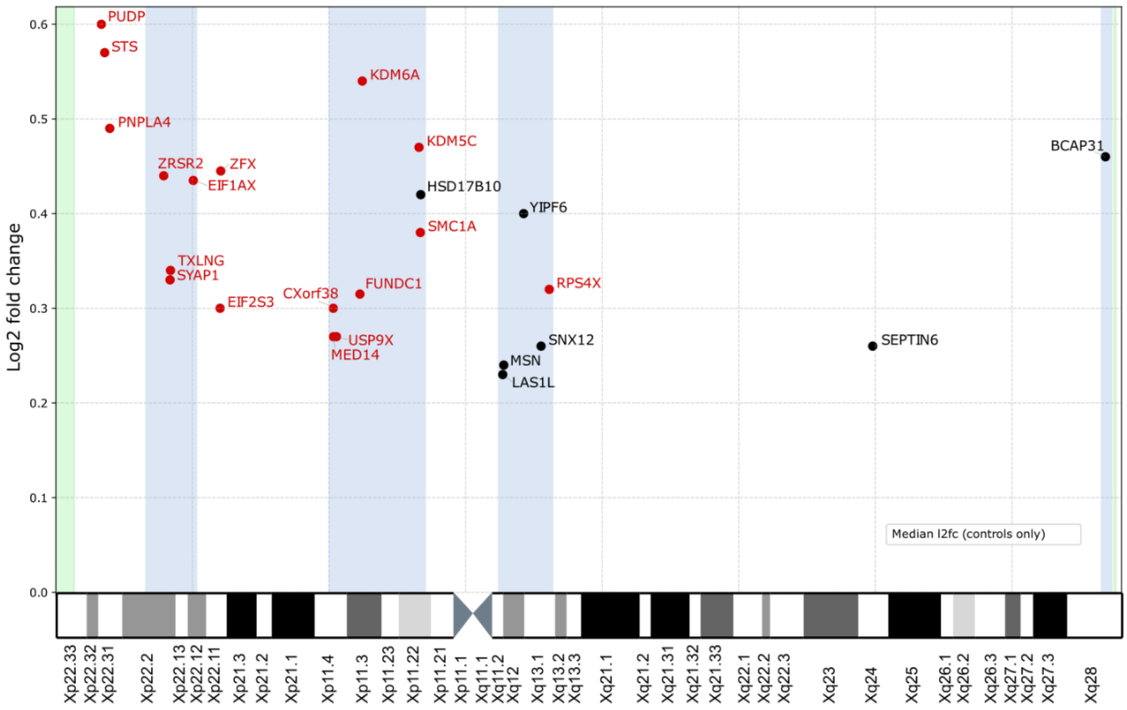


C

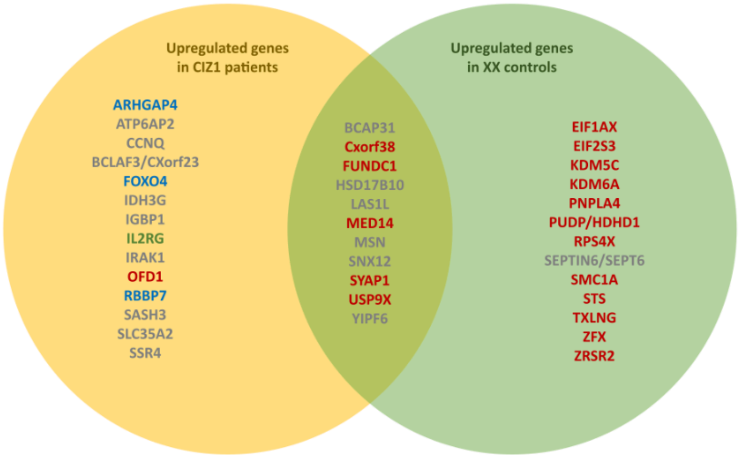
